# Meta-analysis methods for risk difference: a comparison of different models

**DOI:** 10.1101/2022.05.06.22274777

**Authors:** Juanru Guo, Mengli Xiao, Haitao Chu, Lifeng Lin

## Abstract

Risk difference is a frequently-used effect measure for binary outcomes. In a meta-analysis, commonly-used methods to synthesize risk differences include: 1) the two-step methods that estimate study-specific risk differences first, then followed by the univariate common-effect model, fixed-effects model, or random-effects models; and 2) the one-step methods using bivariate random-effects models to estimate the summary risk difference from study-specific risks. These methods are expected to have similar performance when the number of studies is large and the event rate is not rare. However, studies with zero events are common in meta-analyses, and bias may occur with the conventional two-step methods from excluding zero-event studies or using an artificial continuity correction to zero events. In contrast, zero-event studies can be included and modeled by bivariate random-effects models in a single step. This article compares various methods to estimate risk differences in meta-analyses. Specifically, we present two case studies and three simulation studies to compare the performance of conventional two-step methods and bivariate random-effects models in the presence or absence of zero-event studies. In conclusion, we recommend researchers using bivariate random-effects models to estimate risk differences in meta-analyses, particularly in the presence of zero events.

## 1 Introduction

Meta-analysis is a set of statistical tools commonly used in biomedical research and other disciplines to combine and contrast the information from multiple independent studies on the same question, with the aim to synthesize evidence that can be used to underpin guidelines, patient decision aids, and other products that shape health care. ^1^ It could help detect and reduce multiple types of biases, including publication bias and outcome reporting bias, ^2–4^ improve statistical efficiency (over individual studies), and identify patterns and sources of disagreement among different study results. ^5,6^ The growth of interest in evidence-based medicine and comparative effectiveness research has led to a dramatic increase in systematic reviews and meta-analyses, with over 10,000 peer-reviewed publications per year. ^7^

When the outcome is binary, a meta-analysis typically involves the computation of a weighted average of summary statistics based on the 2 × 2 table data from each study. Three effect measures are commonly used: the odds ratio, risk ratio, and risk difference (RD). ^8^ The odds ratio and risk ratio are both multiplicative measures of effect, while the RD is an absolute measure of effect. The selection of effect estimation for meta-analyses of binary outcomes remains much debated in the literature. ^9–15^ The choice of estimation methods becomes more challenging when estimation difficulty occurs, and existing studies have extensively compared various estimation methods for the odds ratio and risk ratio. ^16–18^ However, meta-analysis methods for synthesizing RDs have not been thoroughly compared. Given the importance of the RD in clinical decision-making, ^19^ this article aims to compare different methods of estimating the RDs in a meta-analysis.

Two frequently-used approaches to synthesizing RDs are the two-step methods and one-step methods. The two-step methods first estimate the RD for each study separately, and then summarize those estimates from the first step into the overall RD with study-specific weights. The estimates from the first step are assumed to follow normal distributions. Three models are widely used in the two-step methods: the common-effect (CE), the fixed-effects (FE), or the random-effects (RE) model. ^20^

The CE model assumes the same RD among all studies in a meta-analysis. The FE model makes a less stringent assumption that all studies have different expected RDs, but the studies are not considered as a random sample of all possible studies. The RE model, on the other hand, assumes that the studies included in the meta-analysis represent a random sample of all possible studies, ^21^ and the true RD of each study varies randomly from study to study according to a random distribution (commonly normal without any justification). In the CE model, the RD is estimated mainly by the inverse-variance (IV) method and Mantel–Haenszel (MH) method, where they define different weights according to different criteria. ^22^ The FE model estimates the RD as a population parameter considering the true treatment effect is different for each study. For the RE model, the between-study variation (heterogeneity, *τ*^2^) is first estimated and then is incorporated into the weighted RD estimate. One of the most popular methods to estimate between-study heterogeneity is the DerSimonian–Laird (DL) estimator, ^23^ however, recent research shows that the DL estimator can be biased with incorrectly high precision. ^24^ Many other heterogeneity estimators are available, including Hedges (HE), Paule– Mandel (PM), maximum-likelihood (ML), restricted maximum-likelihood (REML), Hunter– Schmidt (HS), Sidik–Jonkman (SJ), and empirical Bayes (EB) estimators. Each has its own pros and cons. ^23^

All the two-step methods need continuity correction on the zero events when the weights in the second step involve zero event probability. In contrast, the one-step methods model the data from binomial distributions directly such that the parameters are estimated by the ML method in one step. The one-step methods originate from the Sarmanov family of bivariate distributions and mixed-effects model for correlated binary outcomes. The widely-used models for one-step methods are the bivariate random-effects models, including the bivariate generalized mixed-effects model (BGLMM) and bivariate beta-binominal model (BBBM). ^25,26^ In the meta-analysis, they model the events between the treatment and control groups with their potential correlations based on BGLMMs or BBBMs. ^27^

When the events are not rare and no extra corrections are required, the two-step methods perform similarly to one-step methods for the same dataset. ^17^ However, if the probability of the events is rare or if the study sample sizes are small, the normality assumptions in the two-step models might not hold. In addition, zero-event studies lead to a zero variance estimate. When both the treatment and control groups have zero events, i.e., double-zero-events (DZEs), two-step methods cannot define the weight in the IV method. Many solutions were proposed to tackle this problem. ^18^ Either studies with DZEs are not included in the meta-analysis, or they are included after applying the continuity correction; both approaches bias the result. ^28,29^ Such problems would not occur with the one-step method; DZEs are modeled directly by bivariate random-effects models. ^30^

In this article, methods for estimating the RD are discussed in their mathematical formulation and compared empirically. We first describe the statistical rationale of all methods mentioned in Section 2. Then, Section 3 shows the results of methods for two real case studies, one without zero-event studies and one with zero-event studies. In Section 4, we explore and contrast the performance of each method by simulated meta-analyses under a variety of conditions. We will discuss our findings and conclude in Section 5.

## 2 Methods

Consider a meta-analysis of *K* studies with binary outcomes, and let *k* = 1, 2, …, *K* index the studies. For study *k*, we denote the number of patients in the experimental group by *n*_1*k*_, the number of patients in the control group by *n*_2*k*_, the number of events in the experimental group by *e*_1*k*_, and the number of events in the control group by *e*_2*k*_. In addition, the probability of events in the experimental group is *p*_1*k*_ and that in the control group is *p*_2*k*_. For all the *K* studies, the true overall probability of events in the experimental group is denoted by *p*_1_, and that in the control group is denoted by *p*_2_. The number of events are assumed to follow binomial distributions: *e*_1*k*_ *∼* Binomial (*n*_1*k*_, *p*_1*k*_) and *e*_2*k*_ *∼* Binomial (*n*_2*k*_, *p*_2*k*_). The RD measures the difference in the event probabilities between the experimental and control groups, i.e., *δ*_*k*_ = *p*_1*k*_ *− p*_2*k*_ in study *k*. When one assumes studies share the same true RD, *δ*_*k*_ becomes *δ*, which is the true overall RD across all studies. Let *δ*_method_ be *δ* for a particular method, e.g., *δ*_CE_ means *δ* in the CE model. The observed RD in study *k* is denoted by *d*_*k*_.

The two-step methods first estimate the RD of each study and then calculate the pooled RD. In the first step, the probability of events of each group is estimated as 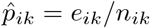 for *i* = 1, 2, and its variance estimate is 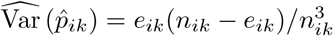. The estimate of the RD for study *k* is 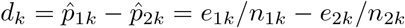 with variance estimate

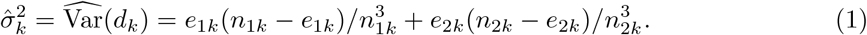

In cases of DZE studies (with *e*_*ik*_ = 0 or *e*_*ik*_ = *n*_*ik*_), it is questionable that the variance estimate turns to be zero. In the second step, the RDs *d*_*k*_’s from the individual studies are pooled together via certain meta-analysis methods such as the CE, FE, and RE models as follows.

### 2.1 Two-step method: common-effect model

One simple meta-analysis approach is to assume a common RD across all studies, i.e., the CE model, which is also commonly referred to as the FE model or equal-effect model. ^22,31^ We will distinguish between the CE model and FE model in Section 2.2. Under the CE model assumption, each component study comes from the same population where the differences between the observed effects are only due to random variations. The model is written as:

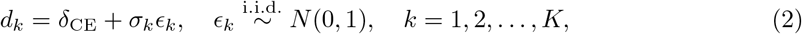

where *ϵ*_*k*_ is the random variation of observed RD from *δ*_CE_, and the *σ*_*k*_ describes the variation. The variance estimate is given by Equation (1). Conventionally, the estimated standard errors 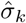 are used to replace the true standard errors *σ*_*k*_ in Equation (2). Because the estimate of 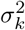 in a DZE study is zero, DZEs may cause estimation problems for *δ*_CE_. In this case, an estimate to *δ*_CE_ needs some adjustments of the observed RDs from individual studies. ^32^

#### 2.1.1 Inverse-variance method

The IV method is one of the classical methods for implementing the CE model. ^33^ It estimates the pooled RD as a weighted average of individual RD of each study, where the weight given to each study is the inverse of the variance of the RD:

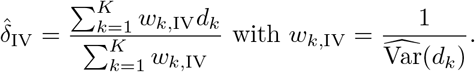

The variance estimates are zero in DZE studies, making the IV estimate incalculable. There are two ways to deal with this problem. One way is to remove the DZE studies. Another way is to add a continuity correction *c*_1*k*_ to *e*_1*k*_ and *n*_1*k*_ *− e*_1*k*_ and add *c*_2*k*_ to *e*_2*k*_ and *n*_2*k*_ *− e*_2*k*_; a common choice of the continuity correction is *c*_1*k*_ = *c*_2*k*_ = 0.5. An alternative continuity correction is *c*_1*k*_ and *c*_2*k*_ that satisfy *c*_1*k*_*/c*_2*k*_ = *n*_1*k*_*/n*_2*k*_ and *c*_1*k*_ + *c*_2*k*_ = 1; we call this correction method as the treatment arm continuity correction (TACC). ^29,34^

#### 2.1.2 Mantel–Haenszel method

The MH method is another popular method for the CE model. ^35,36^ It shares the same idea with the IV method as it estimates the RD by the weighted average of individual RD of each study. The difference is that MH method uses sample sizes to define weights:

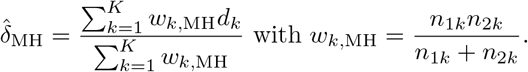

The advantage of this method is that it can naturally handle DZE studies as the weights are based on sample sizes that are always positive, instead of variances. Nevertheless, a continuity correction is still applied to the MH method as a convention. ^29^

### 2.2 Two-step method: fixed-effects model

The CE model assumes that all studies have the same effect, but this assumption may not strictly hold in real-world settings. The FE model permits each study to have different true treatment effects: ^31,37^

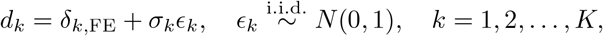

where *δ*_*k*,FE_ denotes the true RD of study *k*. Different types of weights could be given to *d*_*k*_ to estimate *δ*_*k*,FE_. A commonly-used weight is the inverse of the observed sample variance, i.e,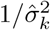. In this case, the estimation of the FE model is the same as the CE model using the IV method. ^20^

### 2.3 Two-step method: random-effects model

In real-world cases, it is common that different component studies come from heterogeneous populations. ^38,39^ Consequently, the true RDs in each study are different, rather than sharing a common RD, e.g., *δ*_IV_. The heterogeneity statistic *Q* can test whether RDs are heterogeneous. ^40^ It is defined as 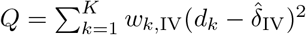 and follows approximately a 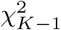 distribution under the null hypothesis of homogeneity, i.e., *H*_0_ : *δ*_*k*_ = *δ* (*k* = 1, 2, …, *K*). If the null hypothesis is rejected, then the effects are different across studies due to their heterogeneous populations. To account for the heterogeneity of study-level true effects, the RE model uses random effects to represent the between-study heterogeneity. The true study effects are assumed to follow a normal distribution with mean zero and a between-study variance *τ*^2^:

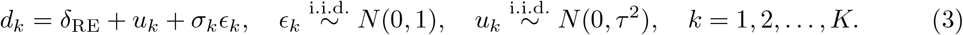

Here, the summarized RD is *δ*_RE_, and *u*_*k*_ is the random effect due to between-study heterogeneity. This model aims to estimate both *δ*_RE_ and *τ*. We can first estimate *τ* and then incorporate *τ* in IV method in the CE model to estimate *δ*_RE_, i.e., 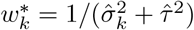. The CE model is a special case of the RE model when *τ* = 0. As in the CE model, the RE model still has technical difficulties in the presence of DZE studies. The RE model may predict a RD outside the range of −1 to 1 due to unbounded normally distributed random effects. An accurate estimate of the between-study variance (*τ*^2^) may reduce such a problem. Many estimators of the between-study variance *τ*^2^ exist ^32,41^; we review 8 of them as follows.

#### 2.3.1 DerSimonian–Laird estimator

The DL estimator is the most widely used method to estimate *τ*^2^ in the current literature. ^42^ The estimator is given by:

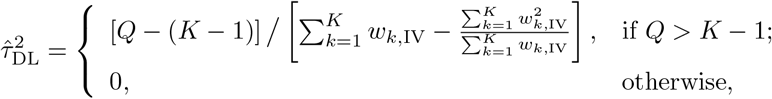

where *Q* is the the heterogeneity statistic.

#### 2.3.2 Hedges estimator

The HE estimator is an unbiased estimator of *τ*^2^ given by: ^43^

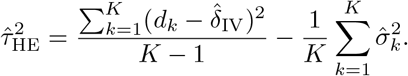

The major advantage of HE estimator is its simplicity because it does not consider different weights across studies. It can be used as an initial value for some estimators that need to be obtained via iterations.

#### 2.3.3 Paule–Mandel estimator

The PM estimator applies iterations to the generalized *Q* statistic: ^44^

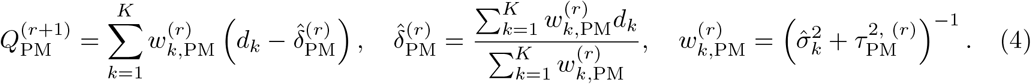

The generalized *Q* statistic is expected to equal *K −* 1, and 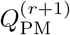 is the generalized *Q* statistic at the (*r* + 1)th iteration. The 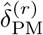 and 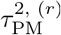 are the estimates of *δ*_PM_ and 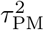 at the *r*th iteration. The iterations start with a value of 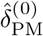 just slightly over zero and stop when the change in 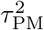 is less than a specific number or Equation (4) satisfies.

#### 2.3.4 Maximum-likelihood estimator

The RE model in Equation (3) is essentially a linear mixed model, which could be implemented by the ML estimation. ^45^ The log-likelihood function which is given by:

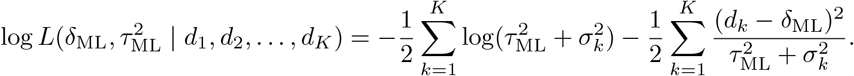

Taking partial derivatives to the log-likelihood function, 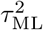 could be solved via the following equation:

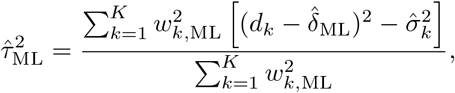

where 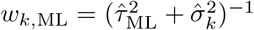 and 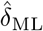 is the estimated *δ*_ML_. We estimate 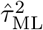 and *δ*_ML_ through iterations; the iterations can start from an initial value of 0 for *δ*_ML_ and use the estimate of *δ* from other methods (e.g., 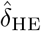) as the initial value for *δ*_ML_. The convergence is typically achieved within 10 iterations. ^46^

#### 2.3.5 Restricted maximum-likelihood estimator

The ML estimator fails to account for the heterogeneity of the data in the first step of iteration and is sometimes negatively biased; the REML estimator solves these problems by a transformation. ^45,47^ Specifically, the REML estimation modifies the log-likelihood function as

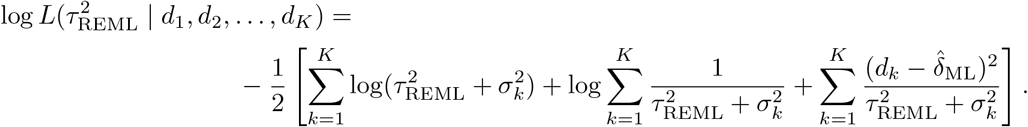

The estimator of 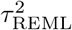 is obtained from solving the following equation:

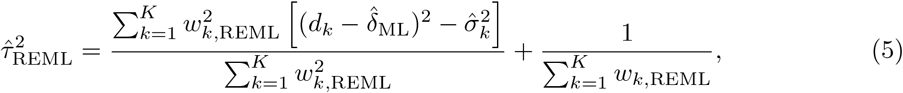

where 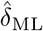 is the likelihood-based estimate of *δ*, which is identical to the one obtained as the ML estimator. The weight *w*_*k*,REML_ is 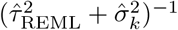. This estimation also requires iterations. Of note, the REML estimator may be obtained using an “approximate” equation: ^48–50^

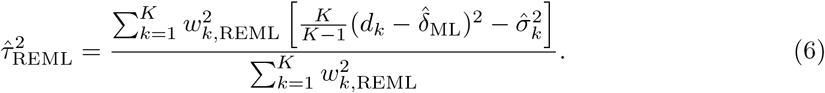

Equations (5) and (6) are equivalent only when studies are homogeneous. This situation is rare in practice, so Equation (5) should be preferred. ^45^

#### 2.3.6 Hunter–Schmidt estimator

The HS estimator is an unbiased estimate of *τ* ^2, 51^ which is given by

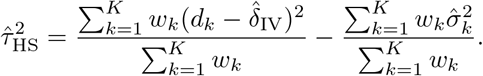

Here, *w*_*k*_ can be any valid weights, and a convenient type of weights is *w*_*k*,IV_.

#### 2.3.7 Sidik–Jonkman estimator

The SJ estimator is a general between-study variance estimator that is based on the weighted residual sum of squares as follows: ^52^

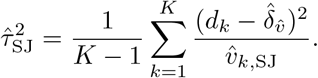

Here, 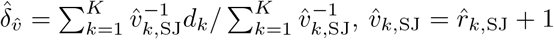, and 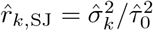, where 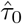 is a rough estimate of *τ* such that 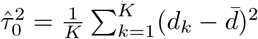 with 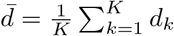.

#### 2.3.8 Empirical Bayes estimator

The EB estimator is a multi-parameter shrinkage estimator of *τ*. ^53^ If *δ*_*k*_ is the true effect size for study *k* and 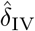 is the effect estimate from the IV method, then

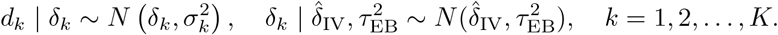

The posterior distribution of *δ*_*k*_ is:

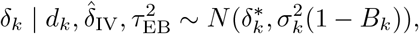

where 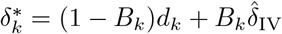 and 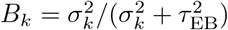. Similar to 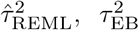 can be estimated by iterations using the following equations if the within-study variances 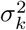 are homogeneous:

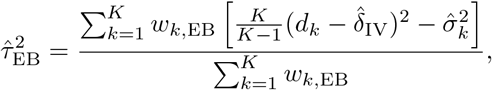

where 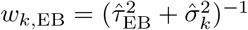. As discussed earlier, 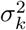 often differs among studies in a meta-analysis. In that case, 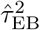 is obtained by the EM algorithm. ^54^ The EB estimator is essentially the same as the PM estimator. ^55^

### 2.4 One-step methods: bivariate random-effects models

Contrary to the two-step methods introduced above, the one-step methods do not force a normal distribution on the estimated RDs from individual studies. Instead, they typically assume that the event counts of individual studies follow binomial distributions. Thus, the one-step methods need no continuity correction for zero counts and avoid bias from continuity corrections. Here, we focus on the bivariate random-effects models:

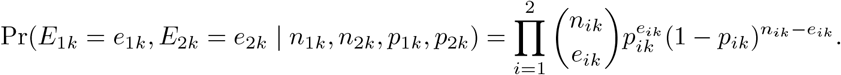

The joint distribution of *p*_1*k*_ and *p*_2*k*_ can be specified across all studies. There are two main model formulations under the bivariate random-effects models for meta-analysis of comparative studies, i.e., the BGLMM and BBBM. ^27^

#### 2.4.1 Bivariate generalized linear mixed-effects model

The BGLMM transforms the binomial distributed probabilities *p*_1*k*_ and *p*_2*k*_ into normal random variables as: ^27^

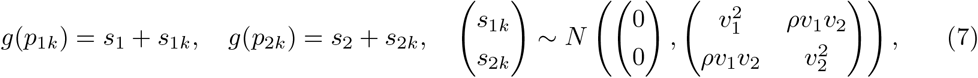

where the link function *g*(·) is usually the logit or probit link. To facilitate the estimation, the inverse of Fisher’s *z* transformation, *ρ* = [exp(2*z*) − 1]*/*[exp(2*z*) + 1], is used to transform the correlation coefficient. ^56^ The parameters *v*_1_, *v*_2_, and *ρ* can be obtained via estimation methods such as the ML. The overall mean of *p*_1*k*_ and *p*_2*k*_ (*k* = 1, 2, …, *K*) across studies, i.e., *p*_1_ and *p*_2_, are calculated as follows:

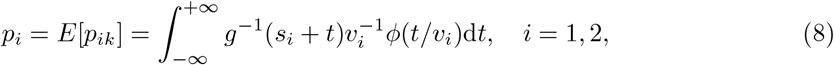

where *ϕ*(·) denotes the standard normal density. Then, the overall RD by the BGLMM is *δ*_BGLMM_ = *p*_1_ *− p*_2_.

#### 2.4.2 Bivariate beta-binomial model

In the BBBM, the joint distribution of *p*_1*k*_ and *p*_2*k*_ is modeled by the Sarmanov bivariate beta distribution in their original scales as: ^27,57^

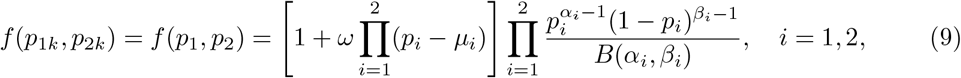

where *α*_*i*_, *β*_*i*_ *>* 0, *ω* is a real number that satisfies 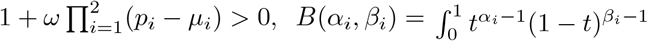d*t* is the beta function, and *µ*_*i*_ is the mean of the binomial event probability, i.e., *E*[*p*_*i*_] = *µ*_*i*_ = *α*_*i*_*/*(*α*_*i*_ + *β*_*i*_). ^58^ The correlation coefficient between the two groups is 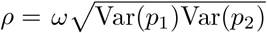, where Var(*p*_*i*_) = *α*_*i*_*β*_*i*_*/*[(*α*_*i*_ + *β*_*i*_)^2^(*α*_*i*_ + *β*_*i*_ + 1)] and *ω* depends on *ρ* and Var(*p*_*i*_). To ensure that *ρ* lies between −1 and 1, we set some constraints on *ω* as follow: ^27^

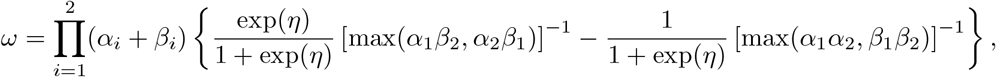

where *η* is a real number.

Consequently, the marginal joint distribution function of the BBBM is

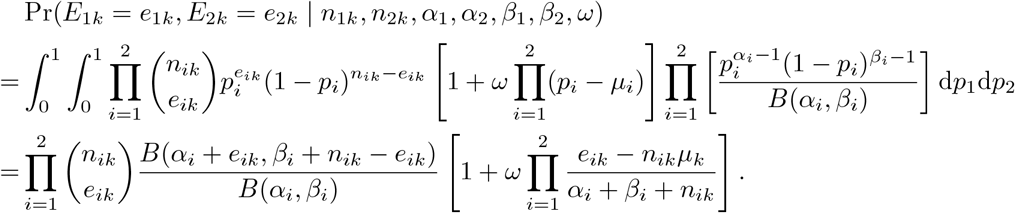

The ML estimation could be used to estimate the parameters in this model and the log-likelihood function is

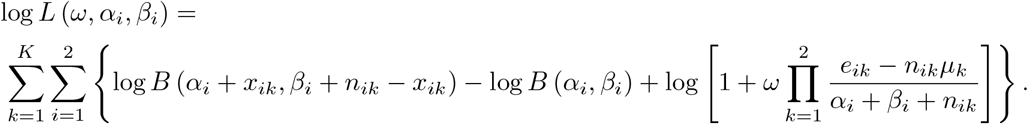

If *α*_*i*_ and *β*_*i*_ are known, the overall RD from the BBBM can be calculated as *δ*_BBBM_ = *E*[*p*_1*k*_] *− E*[*p*_2*k*_] = *α*_1_*/*(*α*_1_ + *β*_1_) *− α*_2_*/*(*α*_2_ + *β*_2_).

### 2.5 Implementation

In this article, all the two-step methods are implemented by the “meta” package (version 5.2-0) in R (version 4.1.3). ^32,59^ The bivariate random-effects models are implemented by SAS (version 9.4) using PROC NLMIXED. ^60^ It uses an adaptive Gaussian quadrature to approximate the likelihood integrated over the random effects, and it maximizes the likelihood function by dual quasi-Newton optimization techniques for the BGLMM and BBBM.

## 3 Case studies

In order to show and compare the empirical performance of various methods in estimating RDs, we apply the above methods to two real datasets, one without zero events and another with zero events, as shown in Tables 1 and 2. Detailed code for case studies could be found in the Supplementary Materials.

**Table 1.** Dataset of risk of stunting in left-behind children by Fellmeth et al. ^61^ (16 studies without zero events).

| Study | Left-behind children |  | Children not left-behind |  |
| --- | --- | --- | --- | --- |
| | $e_1$ | $n_1$ | $e_2$ | $n_2$ |
| Graham and Jordan (2013a) | 22 | 237 | 54 | 243 |
| Graham and Jordan (2013b) | 41 | 255 | 42 | 227 |
| Ban et al. (2017) | 379 | 2426 | 609 | 3708 |
| Feng et al. (2010) | 496 | 1132 | 499 | 1095 |
| Wang et al. (2011) | 25 | 590 | 12 | 285 |
| Mou et al. (2009) | 1276 | 7585 | 1231 | 7557 |
| Xie et al. (2013) | 212 | 675 | 133 | 441 |
| Chen et al. (2013) | 224 | 1157 | 193 | 1157 |
| Gao et al. (2010) | 33 | 541 | 126 | 2445 |
| Onyango et al. (1994) | 33 | 69 | 33 | 85 |
| Xia et al. (2011) | 77 | 318 | 138 | 757 |
| Pan and Chen (2014) | 38 | 309 | 38 | 460 |
| Yu et al. (2013) | 31 | 200 | 204 | 1961 |
| Chen et al. (2012) | 54 | 362 | 12 | 148 |
| Li et al. (2011) | 44 | 738 | 18 | 789 |
| Tao et al. (2016) | 23 | 472 | 3 | 355 |

$e_1$ : the number of events in the experimental group; $n_1$ : the number of patients in the experimental group; $e_2$ : the number of events in the control group; $n_2$ : the number of patients in the control group.

**Table 2.** Dataset of risk of COVID-19 and related diseases with face masks by Chu et al. ^62^ (26 studies containing zero events).

| Study | Face mask |  | No face mask |  |
| --- | --- | --- | --- | --- |
| | $e_1$ | $n_1$ | $e_2$ | $n_2$ |
| Scales et al. (2003) | 3 | 16 | 4 | 15 |
| Liu et al. (2009) | 8 | 123 | 43 | 354 |
| Pei et al. (2006) | 11 | 98 | 61 | 115 |
| Yin et al. (2004) | 46 | 202 | 31 | 55 |
| Park et al. (2016) | 3 | 24 | 2 | 4 |
| Kim et al. (2016)* | 0 | 7 | 1 | 2 |
| Heinzerling et al. (2020)* | 0 | 31 | 3 | 6 |
| Nishiura et al. (2005) | 8 | 43 | 17 | 72 |
| Nishiyama et al. (2008) | 17 | 61 | 14 | 18 |
| Reynolds et al. (2006) | 8 | 42 | 14 | 25 |
| Loeb et al. (2004) | 3 | 23 | 5 | 9 |
| Wang et al. (2020)* | 0 | 278 | 10 | 215 |
| Seto et al. (2003)* | 0 | 51 | 13 | 203 |
| Wang et al. (2020) | 1 | 1286 | 119 | 4036 |
| Alraddadi et al. (2016) | 6 | 116 | 12 | 101 |
| Ho et al. (2004) | 2 | 62 | 2 | 10 |
| Teleman et al. (2004) | 3 | 26 | 33 | 60 |
| Wilder-Smith et al. (2005) | 6 | 27 | 39 | 71 |
| Ki et al. (2019)* | 0 | 218 | 6 | 230 |
| Kim et al. (2016) | 1 | 444 | 16 | 308 |
| Hall et al. (2014)** | 0 | 42 | 0 | 6 |
| Ryu et al. (2019)** | 0 | 24 | 0 | 10 |
| Park et al. (2004)** | 0 | 60 | 0 | 45 |
| Peck et al. (2004)** | 0 | 13 | 0 | 19 |
| Burke et al. (2020)** | 0 | 64 | 0 | 13 |
| Ha et al. (2004)** | 0 | 61 | 0 | 1 |

\*single-zero-event study; \*\*double-zero-event study.
$e_1$ : the number of events in the experimental group; $n_1$ : the number of patients in the experimental group; $e_2$ : the number of events in the control group; $n_2$ : the number of patients in the control group.

### 3.1 Datasets

#### 3.1.1 Risk of stunting in left-behind children

Fellmeth et al. ^61^ presented a meta-analysis investigating whether left-behind children and adolescents had different health conditions compared with children and adolescents with non-migrant parents. We focus on stunting as a binary outcome. This research identified 16 studies without zero events, including 38,779 children (17,066 left-behind children and adolescents and 21,713 children and adolescents with non-migrant parents). Because this dataset contains no zero events, we can use it to investigate whether each method gives a consistent estimate of RDs.

#### 3.1.2 Risk of COVID-19 infection with face masks

Due to the widespread of COVID-19 all over the world, evidence on the effectiveness of prevention measures can be combined to inform decisions. Chu et al. ^62^ performed a meta-analysis on different prevention measures in preventing the transmission of COVID-19 and related diseases. We focus on the meta-analysis that compared the use of face masks with no face masks in healthcare settings. The meta-analysis had 26 studies in total, including 5 single-zero-event studies and 6 DZE studies. The total number of patients was 9,445; 6,003 were in the face mask group, and 3,442 were in the no face mask group. This dataset is used to demonstrate the impact of zero-event studies in a meta-analysis.

### 3.2 Results

Tables 3 and 4 present the estimates of the overall RD 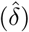and between-study standard deviation 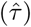 from each model, respectively. All models did not encounter estimation difficulties in the risk of stunting in left-behind children dataset. Similarly, no estimation difficulty occurred in the risk of COVID-19 infection with face masks dataset after corrections for zero events. The implementations of the bivariate random-effects models took slightly longer time than the one-step methods.

**Table 3.** Estimates of the overall risk difference 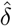 in the two case studies.

|  | Stunting risk<sup>61</sup> | COVID-19 and related diseases risk<sup>62</sup> |  |  |  |
| --- | --- | --- | --- | --- | --- |
| | $\hat{\delta}$ | $\hat{\delta}$ | $\hat{\delta}$ (removing DZE studies) | $\hat{\delta}$ ($c_{1k} = c_{2k} = 0.5$) | $\hat{\delta}$ (TACC) |
| **Common-effect model** |  |  |  |  |  |
| IV method | 0.013 (0.006, 0.020) |  | -0.033 (0.038, -0.029) | -0.032 (-0.037, -0.028) | -0.032 (-0.037, -0.027) |
| MH method | 0.008 (-0.000, 0.015) | -0.071 (-0.080, -0.067) |  |  |  |
| **Random-effects model** |  |  |  |  |  |
| DL estimator | 0.015 (-0.000, 0.030) |  | -0.123 (-0.157, -0.088) | -0.095 (-0.125, -0.065) | -0.089 (-0.118, -0.060) |
| HE estimator | 0.014 (-0.009, 0.037) |  | -0.187 (-0.258, -0.116) | -0.148 (-0.214, -0.082) | -0.144 (-0.211, -0.076) |
| PM estimator | 0.014 (-0.007, 0.036) |  | -0.194 (-0.274, -0.115) | -0.150 (-0.219, -0.081) | -0.144 (-0.212, -0.076) |
| ML estimator | 0.015 (-0.002, 0.032) |  | -0.194 (-0.273, -0.115) | -0.148 (-0.214, -0.082) | -0.141 (-0.205, -0.077) |
| REML estimator | 0.015 (-0.004, 0.033) |  | -0.196 (-0.277, -0.115) | -0.149 (-0.217, -0.081) | -0.142 (-0.209, -0.076) |
| HS estimator | 0.015 (0.001, 0.029) |  | -0.086 (-0.108, -0.063) | -0.070 (-0.091, -0.050) | -0.067 (-0.086, -0.047) |
| SJ estimator | 0.014 (-0.009, 0.037) |  | -0.197 (-0.279, -0.115) | -0.152 (-0.224, -0.080) | -0.146 (-0.218, -0.075) |
| EB estimator | 0.014 (-0.007, 0.036) |  | -0.194 (-0.274, -0.115) | -0.150 (-0.219, -0.081) | -0.144 (-0.212, -0.076) |
| **Bivariate generalized linear mixed-effects model** |  |  |  |  |  |
| probitGLMM | 0.014 (-0.010, 0.038) | -0.153 (-0.209, -0.096) |  |  |  |
| logitGLMM | 0.014 (-0.011, 0.038) | -0.134 (-0.189, -0.079) |  |  |  |
| **Bivariate beta-binomial model** |  |  |  |  |  |
| BBBM | 0.011 (-0.069, 0.091) | -0.164 (-0.264, -0.064) |  |  |  |

DZE: double-zero-event; TACC: treatment arm continuity correction; $c_1$ : continuity correlation of the experimental group; $c_2$ : continuity correlation of the control group.

**Table 4.** Estimates of the between-study standard deviation *τ* from random-effects models in the two case studies.

| Estimator | Stunting risk<sup>61</sup> | COVID-19 and related diseases risk<sup>62</sup> |  |  |
| --- | --- | --- | --- | --- |
| | $\tau$ | $\tau$ (removing DZE studies) | $\tau$ ($c_{1k} = c_{2k} = 0.5$) | $\tau$ (TACC) |
| DL | 0.0231 | 0.0510 | 0.0498 | 0.0487 |
| HE | 0.0409 | 0.1328 | 0.1451 | 0.1531 |
| PM | 0.0373 | 0.1534 | 0.1530 | 0.1543 |
| ML | 0.0274 | 0.1519 | 0.1455 | 0.1439 |
| REML | 0.0301 | 0.1576 | 0.1502 | 0.1486 |
| HS | 0.0209 | 0.0280 | 0.0277 | 0.0276 |
| SJ | 0.0408 | 0.1602 | 0.1624 | 0.1632 |
| EB | 0.0372 | 0.1534 | 0.1530 | 0.1543 |

Table 3 shows that the estimated RDs were more consistent among different models (from 0.008 to 0.015) in the case study of stunting risk than the results in the case study of COVID-19 risk, where different models produced a range of RD estimates from −0.197 to −0.032, likely caused by DZE studies. If DZE studies were removed from the meta-analysis and zero event was not corrected, the RDs were overestimated below 0. Two continuity correction methods, i.e., adding a continuity correction of 0.5 to each cell (*c*_1*k*_ = *c*_2*k*_ = 0.5) and TACC, produced similar estimates. The estimated RDs from bivariate random-effects models were similar or slightly smaller to the estimates from the RE model with continuity corrections. All these estimates suggested a smaller effect of face masks than the RE model with DZE studies removed. Finally, link functions in BGLMMs resulted in little differences in 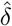.

In the case study of comparing stunting risks between left-behind children and children not left-behind, the *I*^2^ statistic was larger than 70%, ^63^ and the*p*-value of the *Q* test of heterogeneity was less than 0.001. This suggested a high between-study heterogeneity so the RE model was suitable. Different heterogeneity estimators under the RE model gave consistent 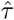 the largest difference between all *τ* estimates was 0.007 (Table 4).

Contrary to the small variation of 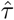 in the case study of stunting risk, a wide range of estimated *τ*^2^ was observed under various heterogeneity estimators in the case study of COVID-19 risk. Because *I*^2^ in this dataset was larger than 80% and the *p*-value of the *Q* test was less than 0.001 in the two-step methods, the RE model was also suitable. Among all the estimators for *τ*, the DL and HS estimators give smaller results than others, leading to substantial differences in 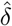.

The considerable differences of the various methods in these case studies leave us to wonder which model provides the best estimates of RDs. Section 4 will use simulated meta-analyses to further investigate the performance of the models.

## 4 Simulation study

### 4.1 Simulation designs

We evaluated different methods for the RD by simulated meta-analyses under different settings using the bivariate probit-normal model. ^64^ Since our main interest is to understand the performance of all models in the presence of DZE, we designed scenarios where heterogeneity is absent or present in a meta-analysis with DZE.

Table 5 outlines settings that simulated three types of meta-analyses: meta-analyses without zero events (Settings 1–4), meta-analyses with zero events and homogeneous studies (Settings 5–8), and meta-analyses with zero events and heterogeneous studies (Settings 9–12). Each meta-analysis type included four simulation settings that were combinations of two different designs of correlations between event probabilities in the treatment and control groups and the number of studies. When no zero events exist in meta-analyses, we used 13 different models introduced in Section 2, including the CE, RE, and bivariate random-effects models. In meta-analyses with zero events, we additionally applied three different continuity corrections to two-step models, resulting in an evaluation of 31 methods in total (Table 6).

**Table 5.** Simulation designs of meta-analyses from a bivariate probit-normal model.

| Setting | $E(p_1)$ | 95% lower<br>confidence limit of $p_1$ | $E(p_2)$ | 95% lower<br>confidence limit of $p_2$ | $\rho$ | $K$ | $n$ |
| --- | --- | --- | --- | --- | --- | --- | --- |
| Meta-analysis without zero events |  |  |  |  |  |  |  |
| 1 | 0.20 | 0.1000 | 0.40 | 0.2000 | 0.3 | 10 | 100 |
| 2 | 0.20 | 0.1000 | 0.40 | 0.2000 | 0.3 | 30 | 100 |
| 3 | 0.20 | 0.1000 | 0.40 | 0.2000 | 0.6 | 10 | 100 |
| 4 | 0.20 | 0.1000 | 0.40 | 0.2000 | 0.6 | 30 | 100 |
| Meta-analysis with zero events and homogeneous studies |  |  |  |  |  |  |  |
| 5 | 0.01 | 0.0050 | 0.03 | 0.0010 | 0.3 | 10 | 100 |
| 6 | 0.01 | 0.0050 | 0.03 | 0.0010 | 0.3 | 30 | 100 |
| 7 | 0.01 | 0.0050 | 0.03 | 0.0010 | 0.6 | 10 | 100 |
| 8 | 0.01 | 0.0050 | 0.03 | 0.0010 | 0.6 | 30 | 100 |
| Meta-analysis with zero events and heterogeneous studies |  |  |  |  |  |  |  |
| 9 | 0.08 | 0.0001 | 0.10 | 0.0005 | 0.3 | 10 | 100 |
| 10 | 0.08 | 0.0001 | 0.10 | 0.0005 | 0.3 | 30 | 100 |
| 11 | 0.08 | 0.0001 | 0.10 | 0.0005 | 0.6 | 10 | 100 |
| 12 | 0.08 | 0.0001 | 0.10 | 0.0005 | 0.6 | 30 | 100 |

**Table 6.**
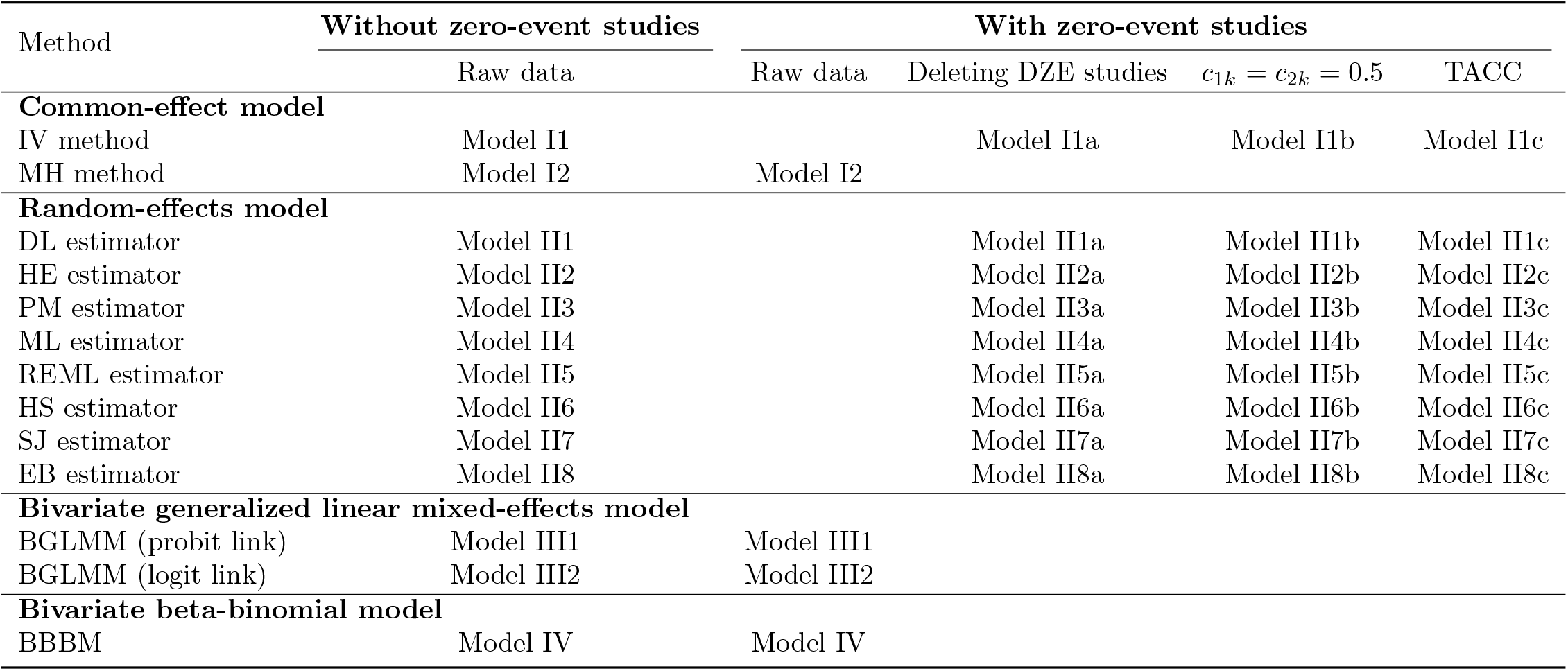
Models in the simulation study for risk differences.

| Method | Without zero-event studies | With zero-event studies |  |  |  |
| --- | --- | --- | --- | --- | --- |
| | Raw data | Raw data | Deleting DZE studies | $c_{1k} = c_{2k} = 0.5$ | TACC |
| **Common-effect model** |  |  |  |  |  |
| IV method | Model I1 |  | Model I1a | Model I1b | Model I1c |
| MH method | Model I2 | Model I2 |  |  |  |
| **Random-effects model** |  |  |  |  |  |
| DL estimator | Model II1 |  | Model II1a | Model II1b | Model II1c |
| HE estimator | Model II2 |  | Model II2a | Model II2b | Model II2c |
| PM estimator | Model II3 |  | Model II3a | Model II3b | Model II3c |
| ML estimator | Model II4 |  | Model II4a | Model II4b | Model II4c |
| REML estimator | Model II5 |  | Model II5a | Model II5b | Model II5c |
| HS estimator | Model II6 |  | Model II6a | Model II6b | Model II6c |
| SJ estimator | Model II7 |  | Model II7a | Model II7b | Model II7c |
| EB estimator | Model II8 |  | Model II8a | Model II8b | Model II8c |
| **Bivariate generalized linear mixed-effects model** |  |  |  |  |  |
| BGLMM (probit link) | Model III1 | Model III1 |  |  |  |
| BGLMM (logit link) | Model III2 | Model III2 |  |  |  |
| **Bivariate beta-binomial model** |  |  |  |  |  |
| BBBM | Model IV | Model IV |  |  |  |

We used bias, mean squared error (MSE), and coverage probability (CP) of nominal 95% confidence interval (CI) of the estimated RD to quantify the methods’ performance. The bias is the difference between an estimate and the true value. The MSE reflects the combination of bias and variance. The CP is the probability that a CI contains the true RD. If an estimate is accurate and precise, we expect that the bias is close to 0, the MSE is small, and the CP is around 95%. ^65^

Under each setting in Table 5, we generated 5,000 meta-analyses. We used the bivariate probit-normal model to generate meta-analyses with binary events, where event counts follow binomial distributions and probit-transformed probabilities follow normal distributions. To be comprehensive in our simulations, we also simulated data from the BBBM; see details in the Supplementary Materials.

In real-world meta-analyses, event probabilities often have within-study correlations between the experimental and control groups. ^66^ We set the correlation coefficient *ρ* to be 0.3 and 0.6. Moreover, we set the number of studies *K* to be 10 and 30. The number of patients in each group for each study (*n*) was 100.

The bivariate probit-normal model includes four other important parameters, i.e., the means and variances of the event probabilities in the probit scale of the two groups. These four parameters affect the likelihood of DZEs occurrence in data. If the event probabilities in both the treatment and control groups are small, then a meta-analysis likely contains DZEs. Nevertheless, simulated meta-analyses with small event probabilities cannot guarantee the presence of zero events, and meta-analyses with large event probabilities may still contain zero events. Therefore, we added further restrictions to the event probabilities by controlling the 95% lower confidence limit of the event probabilities, instead of variances of probabilities, to increase the proportions of meta-analyses with and without zero events under the designed settings. Additionally, we introduced study heterogeneity in a meta-analysis from the difference between the mean and the 95% lower confidence limit of the event probabilities in both groups (*p*_*i*_’s).

We set the mean and 95% lower confidence limit of events for both settings with and without zero events. For meta-analyses without zero events, we set *E*(*p*_1_) = 0.2, *E*(*p*_2_) = 0.4 and set the 95% lower confidence limit of *p*_1_ to 0.1 and 95% lower confidence limit of *p*_1_ to 0.2 across all studies. In meta-analyses with zero events and homogeneous studies, we set *E*(*p*_1_) = 0.01, *E*(*p*_2_) = 0.03 and set the 95% lower confidence limit of *p*_1_ to 0.005 and 95% lower confidence limit of *p*_2_ to 0.01. To introduce heterogeneity in meta-analyses with zero events, we set *E*(*p*_1_) = 0.08 and *E*(*p*_2_) = 0.10 and set the 95% lower confidence limit of *p*_1_ to 0.0001 and 95% lower confidence limit of *p*_2_ to 0.0005. In such cases, the overall *I*^2^ were above 0.80, in contrast to that *I*^2^ *<* 0.20 in settings without heterogeneity. Table 5 gives a summary of simulation designs across all settings.

### 4.2 Simulation results

We summarized the bias (×10^−3^), MSE (×10^−3^), and CP (percentage) of 13 methods in meta-analyses without zero events in Table 7 and those of 31 methods in meta-analyses with zero events in Tables 8 and 9. Figure 1 visualizes the performance comparison among all methods, where each row represents each of the three performance metrics and columns correspond to three meta-analysis types.

**Table 7.** Results of simulated meta-analyses without zero-event studies.

| Method | Setting 1 |  |  | Setting 2 |  |  | Setting 3 |  |  | Setting 4 |  |  |
| --- | --- | --- | --- | --- | --- | --- | --- | --- | --- | --- | --- | --- |
| | Bias<br>($\times 10^{-3}$) | MSE<br>($\times 10^{-3}$) | CP<br>(%) | Bias<br>($\times 10^{-3}$) | MSE<br>($\times 10^{-3}$) | CP<br>(%) | Bias<br>($\times 10^{-3}$) | MSE<br>($\times 10^{-3}$) | CP<br>(%) | Bias<br>($\times 10^{-3}$) | MSE<br>($\times 10^{-3}$) | CP<br>(%) |
| Model I1 | 3.14 | 1.76 | 62.78 | 1.43 | 0.59 | 63.38 | 5.06 | 1.38 | 68.48 | 3.06 | 0.45 | 69.80 |
| Model I2 | 1.15 | 1.64 | 65.08 | -0.62 | 0.54 | 65.66 | 1.10 | 1.26 | 71.16 | -1.64 | 0.41 | 72.76 |
| Model II1 | 1.52 | 1.67 | 90.42 | -0.32 | 0.55 | 93.70 | 2.16 | 1.28 | 90.54 | -0.55 | 0.42 | 93.46 |
| Model II2 | 1.55 | 1.67 | 90.04 | -0.30 | 0.55 | 93.32 | 2.24 | 1.29 | 90.10 | -0.48 | 0.42 | 92.90 |
| Model II3 | 1.54 | 1.67 | 90.16 | -0.31 | 0.55 | 93.46 | 2.20 | 1.28 | 90.42 | -0.50 | 0.42 | 93.22 |
| Model II4 | 1.58 | 1.67 | 88.78 | -0.30 | 0.55 | 92.96 | 2.30 | 1.29 | 88.92 | -0.48 | 0.42 | 92.88 |
| Model II5 | 1.53 | 1.67 | 90.32 | -0.31 | 0.55 | 93.50 | 2.18 | 1.28 | 90.58 | -0.52 | 0.42 | 93.36 |
| Model II6 | 1.57 | 1.67 | 88.94 | -0.31 | 0.55 | 93.18 | 2.29 | 1.29 | 89.02 | -0.50 | 0.42 | 93.08 |
| Model II7 | 1.51 | 1.67 | 91.78 | -0.32 | 0.55 | 94.12 | 2.09 | 1.28 | 92.10 | -0.60 | 0.42 | 94.32 |
| Model II8 | 1.54 | 1.67 | 90.16 | -0.31 | 0.55 | 93.48 | 2.20 | 1.28 | 90.42 | -0.50 | 0.42 | 93.24 |
| Model III1 | -0.45 | 1.60 | 83.76 | 0.32 | 0.52 | 87.44 | 0.47 | 1.18 | 90.48 | 0.70 | 0.39 | 93.06 |
| Model III2 | -0.11 | 1.61 | 83.92 | -0.37 | 0.52 | 79.56 | -0.17 | 1.18 | 82.11 | -0.03 | 0.40 | 85.14 |
| Model IV | -0.86 | 1.66 | 73.13 | 0.44 | 0.57 | 90.56 | 0.32 | 1.18 | 85.48 | 0.88 | 0.44 | 93.90 |

**Table 8.**
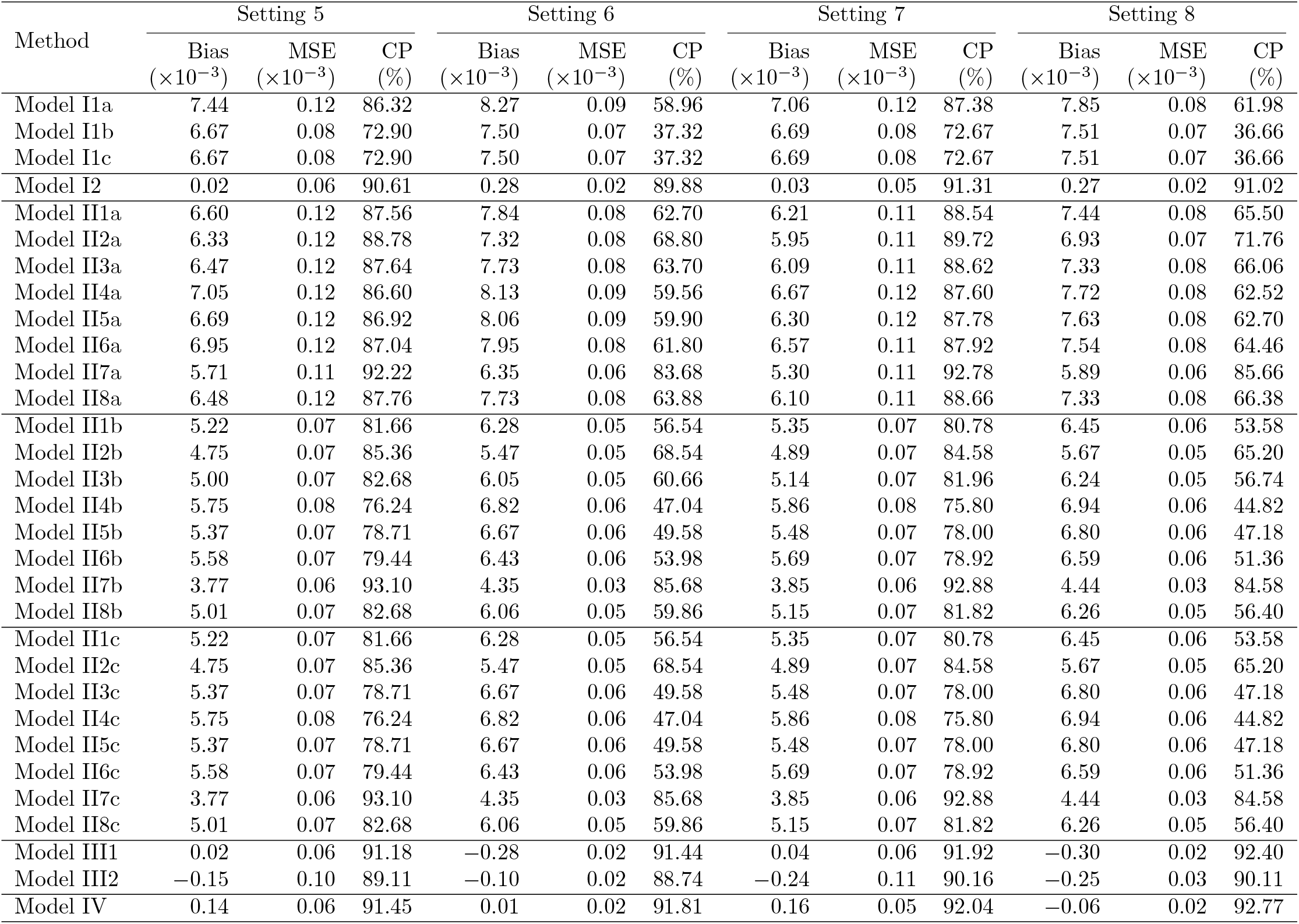
Results of simulated meta-analyses with homogeneous zero-event studies (*I*^2^ *<*0.20).

**Table 9.**
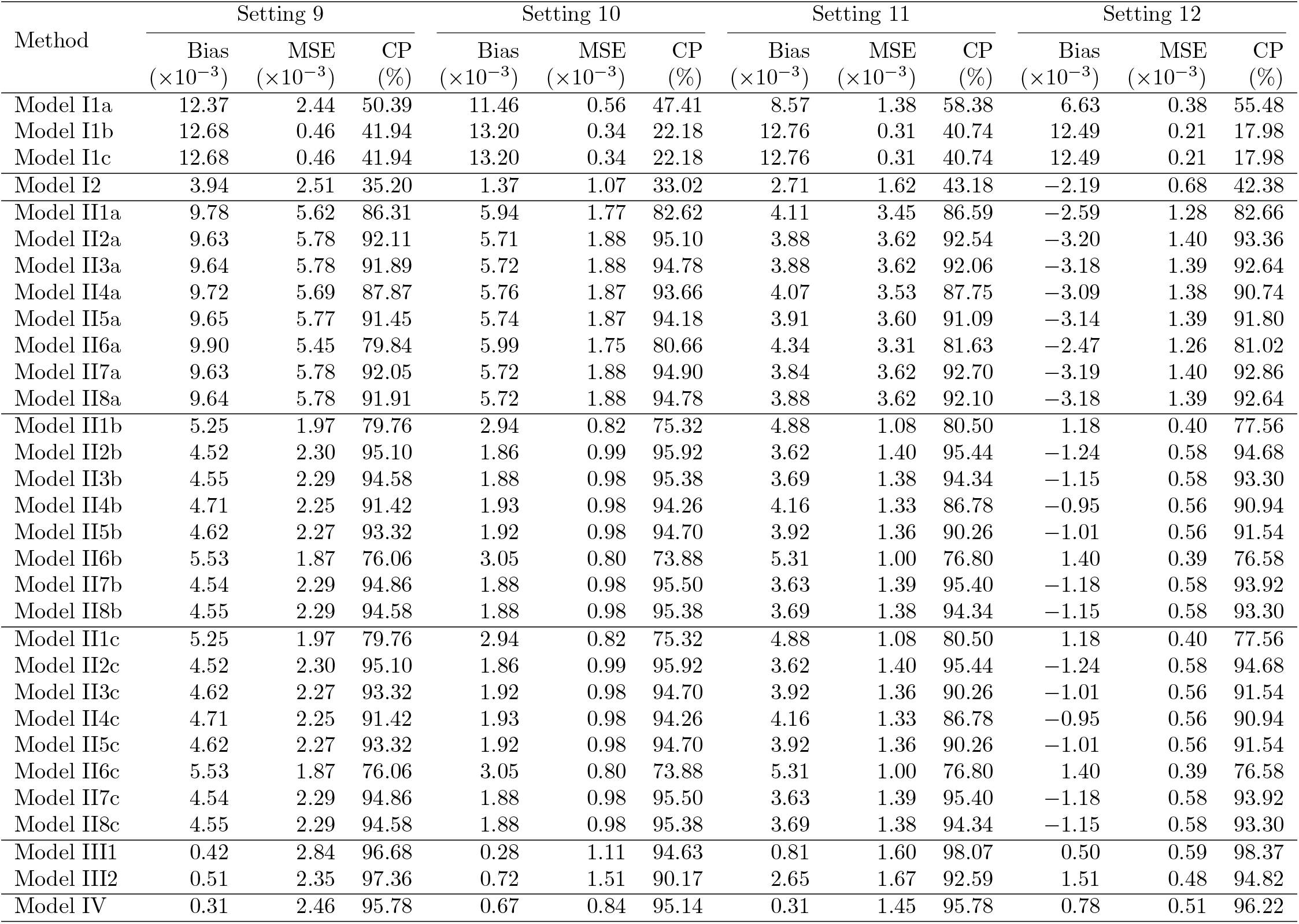
Results of simulated meta-analyses with heterogeneous zero-event studies (*I*^2^ *>*0.80).

**Figure 1.**
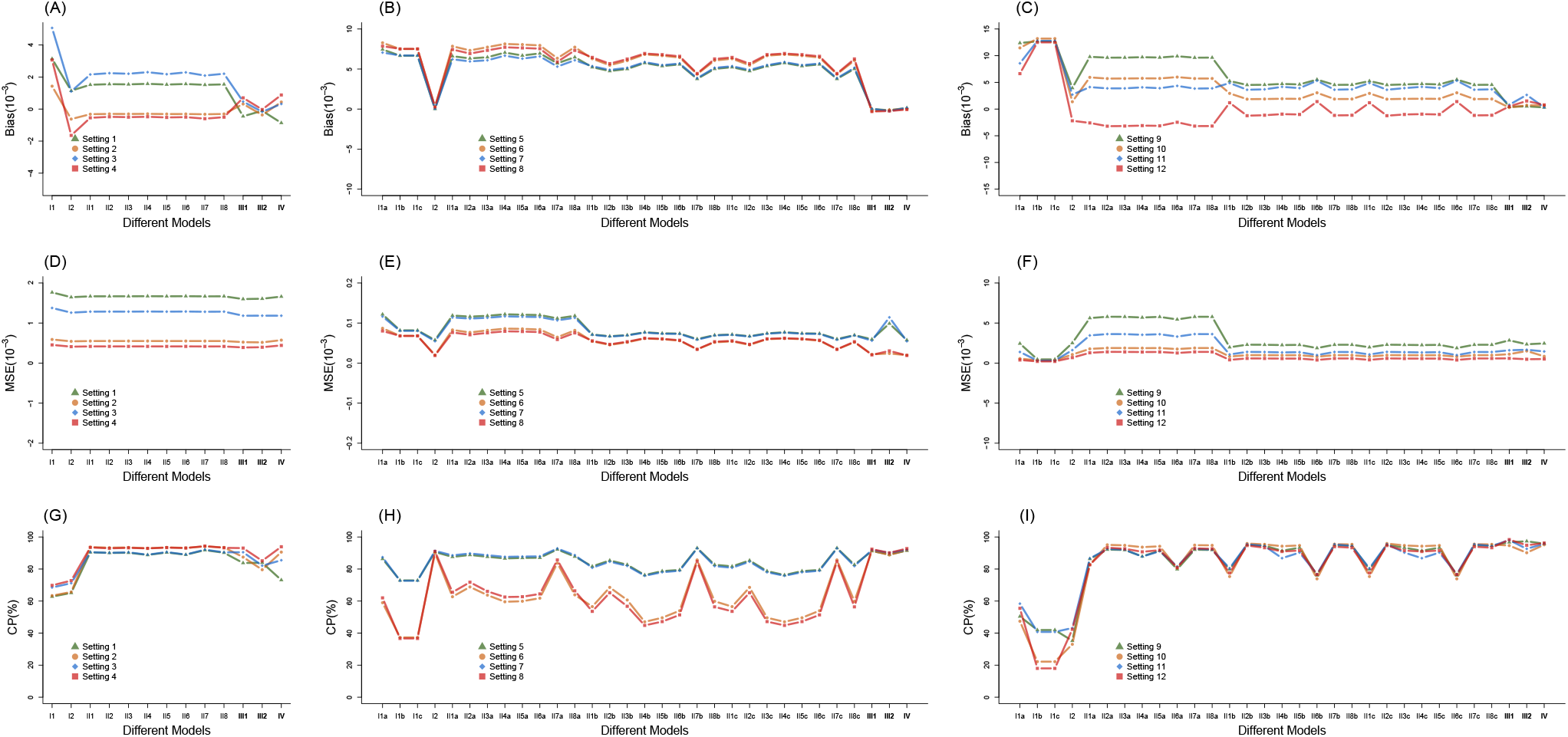
Simulation results for risk differences by various models under different settings.

When the true RD was −0.20 in meta-analyses without zero events, almost every method (except for Models I1 and I2) achieved nearly 90% CP (nominal level of 0.05). While the largest bias was around 0.05 (Model I1), most methods only had a bias of around 0.001. We observed small differences in MSE among all methods. CPs, however, varied across methods; the CE models (Models I1 and I2), in particular, had lower CPs (*<*75%) than other models. The CPs of BBBMs varied from 73.13% to 93.90% under different settings.

In meta-analyses with homogeneous studies and zero events, the performance across models had a greater variation than the previous settings without zero events (Table 8 and Figure 1). The bias ranged from 0.02 × 10^−3^ to 8.27 × 10^−3^. Except for the MH method (Model I2), all two-step models (Model I1a–Model I1c and Model II1a–II8c) had a higher bias than the one-step models (Table 8 and Figure 1). In other words, the two-step models with the continuous correction for zero counts were generally biased. The problem was especially pronounced when a small bias towards the null might lead to an opposite sign to a small true RD. Different methods showed similar MSEs, and the CP ranged from 36.66% to 92.88%. While the one-step model and MH method had a CP *>*85%, two-step models showed a large variation of CP across different choices of models (CE or RE models) or between-study variance estimators in RE models. The variation was greater when the correlation (*ρ*) between treatment and control groups was larger (Settings 7 and 8 in Figure 1H).

Table 9 and Figure 1C, 1F, and 1G showed that one-step methods generally had a better performance than two-step methods when meta-analyses had zero events heterogeneity. The highest average bias was 12.68×10^−3^ in CE model with artificial corrections given the true RD was only 20×10^−3^. Furthermore, most two-step models had higher biases than one-step models, and the bias for two-step models varied by both *ρ* and *K*. Different methods had similar MSEs to each other except for two-step models deleting DZE studies. The CP of all CE models were below 60%, suggesting that CE models were not suitable for meta-analysis with high between-study heterogeneity and zero events. CP for RE models were all larger than 75% and CP for one-step models are all larger than 90%. While BGLMM with the probit link (Mode III1) and BBBM (Model IV) had a consistently low bias and CP of at least 95%, bias or CP in BGLMM with the logit link (Model III2) was sub-optimal when *ρ* was 0.6 in Settings 10 and 12. A possible reason for such a variation is that the logit link is more likely to suffer from estimation difficulties when *ρ* is high in bivariate settings. ^67^ In summary, one-step models, especially BGLMM (probit link) and BBBM, outperformed other models considering all three metrics; thus, they are recommended for estimating meta-analysis with substantial heterogeneity and zero events.

By all evaluation metrics, bivariate random-effects models (Models III1, III2, and IV) had significantly better performance than the others when studies had zero events. Although the CE model with the MH method (Model I2) performed better than most two-step methods when the between-study heterogeneity was low, it had a significantly reduced performance in meta-analyses with high between-study heterogeneity. CE models with the IV method (Models I1a, I1b, and I1c) had poor performance when DZE studies were present, regardless of study heterogeneity. Models that removed DZE studies (Models II1a to II8a) or added continuity corrections (Models II1b to II8b and Models II1c to II8c) not only biased the RD estimate but also increased the MSE and reduced the CP. For a specific model (e.g., Model II4b/II4c vs. Model II4a), the performance of using continuity corrections was slightly better than that of deleting DZE studies. Adding 0.5 to zero events and TACC in DZE had similar performance (Table 8). The variance estimator that produced the best CP depended on specific settings of meta-analyses with DZE. Although two-step models appeared to have similarly large biases (*>* 2 × 10^−3^) for all meta-analyses with DZEs and homogeneous studies (Table 8), only settings with 30 studies (Settings 6 and 8) per meta-analysis had a small CP. This was because a large number of studies (e.g., 30) increased the precision of the RD estimate, and 95% CIs around the biased estimates were unlikely to cover the true RD. This means that most two-step models (except for the CE model with the MH method) were biased in both the point and interval estimate when DZEs studies were present in a meta-analysis with low heterogeneity and a large number of studies. When there was substantial heterogeneity in a meta-analysis with DZEs, bias in all CE models (including the MH method) caused a poor CP.

## 5 Discussion

This article comprehensively evaluates both the two-step and one-step models for estimating the RD in a meta-analysis with binary outcomes. While many existing studies have compared methods in estimating the odds ratio and the risk ratio under various situations, their conclusions may not easily extend to the RD because the RD can be defined in DZEs. In this study, we compared and evaluated multiple models to estimate the RD in meta-analyses with zero-event studies. Results from real-data case studies and simulations show that various models performed consistently in the absence of DZEs but had noticeable differences when DZEs appeared. For datasets with zero-event studies, we suggest using one-step methods, especially BGLMM (probit link) and BBBM. The CE model with the MH method is suitable when zero-event studies are not heterogeneous, in addition to one-step methods. In contrast to other models, these two methods do not need any artificial corrections for zero-event studies. Both had smaller biases and MSEs and higher CPs than other methods. Their performance remains superior under different settings. However, the CE model with the MH method assumes that all studies estimate the same effect, which may not be suitable in the presence of substantial heterogeneity. ^22^ Thus, when studies are heterogeneous, researchers should avoid using the MH method. One-step methods may be preferred over the MH method because they make no explicit assumption about homogeneity.

Although bivariate random-effects models have many advantages, some limitations still remain. Because bivariate random-effects models need sufficient data to estimate model parameters (*p* = 5 in bivariate models), the estimation may encounter singularity problems when the number of studies *n* is small in a meta-analysis. In order to avoid model singularity, the dataset should contain at least six studies such that *n > p*. We suggest researchers use two-step methods instead of bivariate random-effects models if there is a limited number of studies (e.g., *≤*5). In addition, more parameters imply that estimation failures are more likely to arise with inappropriate starting values of parameters in bivariate random-effects models. Finally, some refined CIs of univariate random-effects models exist in the current literature. ^68–70^ While they were not fully investigated in this article, these alternative CI constructions are expected to improve the CPs for two-step methods. Methods to improve the CPs for the bivariate one-step methods demand further research.

Our work may spark future interests in estimating the RD from datasets of 2 × 2 tables, yet several limitations exist. Several RD estimators are not included in this article, such as Bayesian methods. ^29,71^ Bayesian methods have become popular in meta-analyses as they could overcome some computational difficulties encountered by traditional methods and permit the use of pertinent external information for improving the estimation. Current software packages are available to implement our one-step models from Bayesian approaches, such as the PROCBGLIMM in SAS. ^72,73^ Future studies may compare the performance of Bayesian methods with the methods discussed in this article. In addition, one may use other types of bivariate random-effects models, such as the marginal beta-binomial model approach. ^74^ Finally, although we show that one-step models have better performance than two-step models, two-step models that apply artificial corrections currently dominate meta-analysis applications, partly because they are readily implemented by most meta-analysis software. It is worthwhile to develop user-friendly software or modules for bivariate random-effects models for meta-analyses of RDs so that they can be conveniently implemented by practitioners.

## Data Availability

The case study datasets are in Table 1 and 2.

## Supplementary Materials

The Supplementary Materials contain additional simulation studies and statistical code for reproducing the case studies.

## Acknowledgements

Research reported in this publication was partially supported by the National Center for Advancing Translational Sciences grant UL1 TR002494 (HC), the National Library of Medicine grant R01 LM012982 (MX, LL, and HC), and the National Institute of Mental Health grant R03 MH128727 (LL) of the US National Institutes of Health. The content is solely the responsibility of the authors and does not necessarily represent the official views of the National Institutes of Health.

